# Incidence and Case-Fatality Ratio of COVID-19 infection in relation to Tobacco Smoking, Population Density and Age Demographics in the USA: could Particulate Matter derived from Tobacco Smoking act as a Vector for COVID-19 transmission?

**DOI:** 10.1101/2020.10.04.20206383

**Authors:** Yves Muscat Baron

## Abstract

**BACKGROUND:** Tobacco smoking is known to increase the risk for bacterial and viral respiratory infections and this also applies to second-hand smoking. Smoking has been shown to increase the severity of COVID-19 infection and the consequent risk for intra-tracheal ventilation in smokers. Tobacco smoking exposes the user and nearby individuals to very high concentrations of particulate matter in a short period of time. Genes appertaining to COVID-19 have been found adherent to particulate matter. Particulate matter has been shown to travel beyond the social distance of 2 metres up to 10 metres. COVID-19 related mortality has been linked to elevated atmospheric levels of the particulate matter, PM2.5. The aim of the study was to observe the incidence of infection rate and case fatality ratios in the USA, comparing States with partial bans on tobacco smoking, to States with more restrictive smoking regulation, exploring a possible link between smoke-related particulate matter and COVID-19 transmission.

**METHODOLOGY:** Two groups of USA States, differentiated by the degree of smoking legislative restrictions, had a number of variables compared. The incidence of COVID-19 infection, case-fatality ratio and testing frequency were obtained from the John Hopkins Coronavirus Resource Centre. The degree of smoking bans in the USA States was obtained from the websites of the Nonsmokers Rights Foundation. The percentage of the State population which smokes was collected from the Centres of Disease Control database. Population density, Body Mass Index and population percentages of individuals 65+/75+years were obtained from databases concerning USA demographics.

**RESULTS:** With the available data there was no significant difference in COVID-19 testing prevalence between the partial smoking ban group and the more restrictive regulated group. The incidence of COVID-19 infection in the States with limited bans on tobacco smoking was 2046/100,000 (sd+/-827) while the infection incidence in States with more restrictive rulings on tobacco smoking was 1660/100,000 (sd+/-686) (p<0.038). The population percentage of smokers in States with minor limitations to smoking was 18.3% (sd+/-3.28), while States with greater smoking restrictions had a smoking population percentage of 15.2% (sd+/-2.68) (p<0.0006).

The two populations of both groups did not differ numerically (p<0.24) and numbered 157,820,000 in the partial smoking ban group and 161,439,356 in the more restrictive group. Population density correlated significantly with the case-fatality ratio (R=0.66 p<0.0001), as did the 75+year age group (R=0.29 p<0.04). Reflecting the possibility of trans-border transmission, the smoking status of adjacent partial smoking ban States may influence the COVID-19 incidence of bordering States (e.g. Utah) even if the smoking regulations of the latter were stricter than the former.

Other factors that could impact the COVID-19 pandemic in the USA such as the State case-fatality ratio, population density, population percentage with elevated body mass index and the percentage of the state population aged 65years or above did not show any significant difference between both groups of States.

**CONCLUSION:** States in the USA with high levels of tobacco smoking and limited regulation had significantly higher rates of COVID-19 infection incidences than States with greater smoking restrictions. Population density and the age group of 75+years, showed a positive significant correlation with the case-fatality ratio. Besides the adverse effects of tobacco smoking on pulmonary defences, it would be interesting to explore the possibility of infection transmission via coronavirus-laden particulate matter from exhaled fumes derived from tobacco smoking.

## INTRODUCTION

Tobacco smoking is responsible for a significant proportion of global mortality. Of the global burden of 8 million deaths per year, more that 85% of these deaths occur in smokers themselves. Nonsmokers are also at risk, resulting in approximately one million deaths per annum due to second-hand smoking. Second hand smoking exposes bystanders to more than 7,000 chemicals, including about 70 known and probable carcinogens, including toxic and irritant agents. Second-hand smoking exposes these individuals to particulate matter and potentially to respiratory disease through smoke-induced infection transmission (Lubich et al 2011) (U.S. Department of Health and Human Services 2017).

In an effort to curb smoking and reduce the deleterious effects of second-hand smoking, several countries have introduced restrictive measures. In the USA there appears to be a divide, with some States applying restrictive smoking regulation while in others there exist partial smoking bans.

Smoking is associated with significantly elevated human exposure to particulate matter. Long-term exposure to particulate matter PM2.5 in the USA has been linked to COVID-19 related mortality (Wu et al 2020). Particulate matter has been noted to have genes appertaining COVID-17 adherent to it (Setti et al 2020). The possibility of a dual effect of reducing pulmonary defences and the carriage of COVID-19 by particulate matter has been explored by Comunian et al 2020.

This paper assesses whether smoking in USA varies across its member States some of which have less restrictive regulation on tobacco smoking than others. Concomitantly the incidence of COVID-19 across the USA was assessed in relation to the USA’s States reviewed in an effort to find an incidence pattern and case-fatality ratio coinciding with COVID-19 infection. As a corollary the population density body mass index and age demographics were also assessed between both groups of States.

## METHODOLOGY

This study compared two groups of States in the USA, differentiated by the gradation of legislative restrictions on tobacco smoking. The variables assessed include COVID-19 incidence, case-fatality ratios, state population density and population percentages of individuals 65/75years and older. The incidence of COVID-19 infection, prevalence of testing and case-fatality ratios were attained from the John Hopkins Resource Centre database. The degree of smoking restrictions in the States assessed was obtained from the websites of the Nonsmokers’ Rights Foundation. The legislative measures included in the study governed smoking related legal restrictions up to October 2018. The percentage of the state smoking was retrieved from the U.S. Centres of Disease Control database. Population density, body mass index and population percentages of individuals 65/75years and older were obtained from websites concerning USA demographics in 2018 and 2019.

## STATISTICS

The data was analysed for normality by the Smirnov-Kolmogroff test. The percentage of smokers in the state population, incidences of COVID-19 infection, age demographics and body mass index all showed a normal distribution while all the other data assessed were nonparametric. The independent TTest was applied to the population percentage of smokers and incidences of COVID-19 infection. The Mann Whitney U test was applied for comparing nonparametric variables of both groups of States. Pearson correlation was applied for parametric variables and Spearman rank was implemented for nonparametric findings.

## RESULTS

COVID-19 infection incidences were significantly higher in the States with partial bans on tobacco smoking compared with highly regulated States (p<0.038). The incidence of COVID-19 infection in the States with partial bans on tobacco smoking was 2046/100,000 (sd+/-827), while the infection incidence in States with restrictive regulation on tobacco smoking was 1660/100,000 (sd+/-686) (Tables 1. and 2.).

**Table 1.**
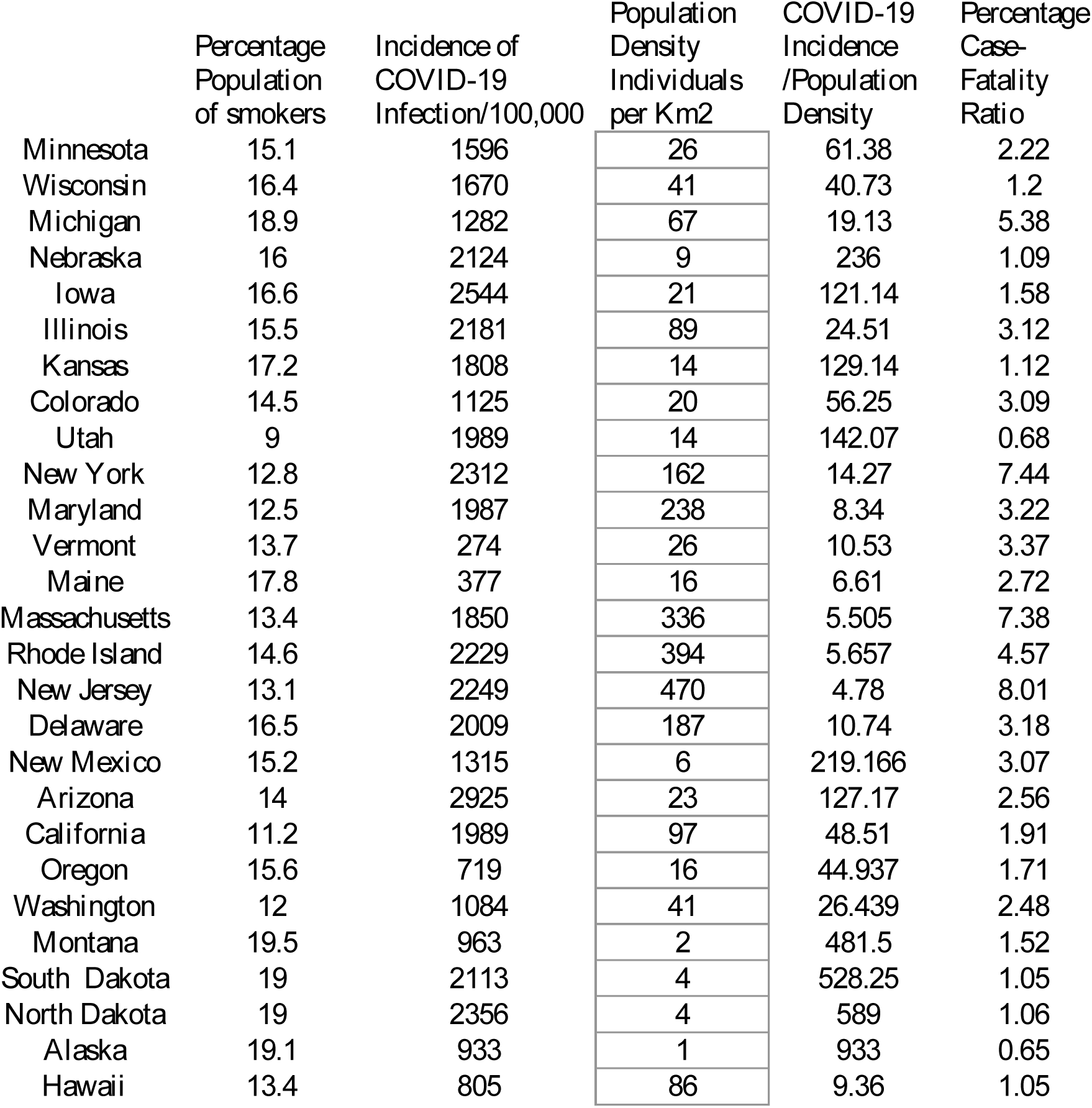
Variables assessed of States with *strict* smoking bans and *lower* smoking population rates

**Table 2.** More variables assessed of States with *strict* smoking regulation and *lower* smoking population rates.

|  | Percentage<br>Population<br>Age >65yr | Percentage<br>Population<br>Age>75yrs<br>Native<br>Born | Percentage<br>Population<br>Age>75yrs<br>Foreign<br>Born | Total<br>Percentage<br>Population<br>Age >75yrs | Percentage<br>of<br>Population<br>Overweight<br>or Obese |
| --- | --- | --- | --- | --- | --- |
| **<u>Table 2.</u>** |  |  |  |  |  |
| Minnesota | 16.3 | 6.38 | 0.35 | 6.73 | 67.3 |
| Wisconsin | 17.5 | 6.82 | 0.32 | 7.14 | 68.4 |
| Michigan | 17.6 | 6.62 | 0.49 | 7.11 | 68.2 |
| Nebraska | 16.1 | 6.58 | 0.22 | 6.8 | 73.7 |
| Iowa | 17.5 | 7.4 | 0.16 | 7.56 | 72 |
| Illinois | 16.1 | 5.73 | 0.92 | 6.65 | 65.8 |
| Kansas | 16.3 | 6.58 | 0.23 | 6.81 | 70.5 |
| Colorado | 15.6 | 4.91 | 0.45 | 5.36 | 61.9 |
| Utah | 11 | 4.12 | 0.28 | 4.4 | 65.2 |
| New York | 16.9 | 5.25 | 1.92 | 7.17 | 57.2 |
| Maryland | 15.9 | 5.53 | 0.84 | 6.37 | 66.5 |
| Vermont | 20 | 7.12 | 0.56 | 7.68 | 66.9 |
| Maine | 21.2 | 8 | 0.38 | 8.38 | 66.4 |
| Massachusetts | 16.8 | 5.77 | 1.2 | 6.97 | 62.1 |
| Rhode Island | 17.7 | 6.56 | 0.94 | 7.5 | 64.4 |
| New Jersey | 16.6 | 5.48 | 1.55 | 7.03 | 65.9 |
| Delaware | 19.4 | 7.03 | 0.5 | 7.53 | 65.9 |
| New Mexico | 18 | 6.44 | 0.59 | 7.03 | 62.8 |
| Arizona | 18 | 6.54 | 0.86 | 7.4 | 66.3 |
| California | 14.8 | 3.93 | 2.1 | 6.03 | 58.5 |
| Oregon | 18 | 6.35 | 0.61 | 6.96 | 64.6 |
| Washington | 15.9 | 5.16 | 0.86 | 6.02 | 63.7 |
| Montana | 19.3 | 7.21 | 0.24 | 7.45 | 60.1 |
| South Dakota | 17.1 | 6.88 | 0.07 | 6.95 | 72.4 |
| North Dakota | 15.8 | 6.81 | 0.09 | 6.9 | 76.9 |
| Alaska | 12 | 3.38 | 0.47 | 3.85 | 72.9 |
| Hawaii | 18.9 | 5.98 | 1.91 | 7.89 | 72.9 |

The percentage of the smoking population in the States with minimum regulation in 2018 was significantly higher that that of States with more severe prohibitions on tobacco smoking (p<0.0006). The percentage of smokers in States with minimal restrictions to smoking was 18.3% (sd+/-3.28) while States with greater smoking restrictions had a smoking population percentage of 15.2% (sd+/-2.68). Other factors that could affect the COVID-19 USA pandemic such as the state case-fatality ratio, population density, body mass index and the percentage of the state population aged 65years or above did not show any significant difference between both groups of States(Tables 1. and 2.).

A number of significant correlations were obtained when comparing variables of all the US States together and when the two groups of States were assessed separately. The States’ population density correlated significantly (R=0.66 p<0.0001) with the case-fatality ratio. The correlation of the population density/case fatality ratio still applied when the two groups of States were assessed separately (partial ban R=0.58 p<0.003 and restrictive regulation R=0.74 p<0.0001) (Tables 1. and 2.).

The case fatality ratio correlated with increasing age in the 75years and over age group (R=0.29 p<0.04). No significant correlation between the 65years and over age group and the case fatality ratio. In the 75 years and over age group the proportion of foreign born population (0.7% sd+/-0.57) was significantly higher in the States with restrictive smoking regulations compared to the population of States with limited smoking restrictions (0.45% sd+/-0.45). There was no difference when comparing the proportion of native born to the foreign born cohorts when the both groups of States were combined in both the 75+ year age group. The correlation between the percentage smoking population and case-fatality ratio just missed statistical significance (R=0.27 p<0.056). There was no significant correlation between the incidence of COVID-19 and case-fatality ratio (Tables 3. and 4.).

**Table 3.** Variables assessed of States with *partial* smoking bans and *higher* smoking population rates.

| **<u>Table 3</u>** | Percentage<br>Population of<br>smokers | Incidence of<br>COVID-19<br>Infection/100,000 | Population<br>Density<br>Individuals<br>per Km2 | COVID-19<br>Incidence<br>/Population<br>Density |
| --- | --- | --- | --- | --- |
| Indiana | 21.1 | 1656 | 71 | 23.32 |
| Missouri | 19.4 | 1860 | 34 | 54.7 |
| Oklahoma | 19.7 | 1941 | 22 | 88.22 |
| Arkansas | 22.7 | 2509 | 22 | 114.04 |
| Tennessee | 20.7 | 2687 | 61 | 44.04 |
| Kentucky | 23.4 | 1377 | 43 | 32.02 |
| Ohio | 20.5 | 1234 | 109 | 11.32 |
| Pennsylvania | 17 | 1210 | 110 | 11 |
| New<br>Hampshire | 15.6 | 584 | 57 | 10.24 |
| Connecticut | 12.3 | 1557 | 286 | 5.44 |
| Virginia | 14.9 | 1644 | 81 | 20.29 |
| North<br>Carolina | 17.4 | 1845 | 79 | 23.35 |
| South<br>Carolina | 18 | 2687 | 62 | 43.338 |
| Georgia | 16.1 | 2883 | 68 | 42.397 |
| Florida | 14.5 | 3183 | 145 | 21.95 |
| Alabama | 19.2 | 2956 | 37 | 79.89 |
| Mississippi | 20.5 | 3137 | 24 | 130.708 |
| Louisiana | 20.5 | 3467 | 41 | 84.56 |
| Texas | 14.4 | 2458 | 40 | 61.45 |
| Nevada | 15.7 | 2461 | 10 | 246.1 |
| Idaho | 14.7 | 2097 | 7 | 299.57 |
| Wyoming | 18 | 841 | 2 | 420.5 |
| West Virginia | 25.2 | 785 | 29 | 481.5 |

**Table 4.** More variables assessed of States with *partial* smoking bans and *higher* smoking population rates.

|  | Percentage<br>Population<br>Age >65yr | Percentage<br>Population<br>Age>75yr<br>Native<br>Born | Percentage<br>Population<br>Age>75yr<br>Foreign<br>Born | Total<br>Percentage<br>Population<br>Age >75yr | Percentage<br>of<br>Population<br>Overweight<br>or Obese |
| --- | --- | --- | --- | --- | --- |
| ***Table 4*** |  |  |  |  |  |
| Indiana | 16.1 | 6.32 | 0.22 | 6.54 | 68.4 |
| Missouri | 17.3 | 6.97 | 0.19 | 7.16 | 68.5 |
| Oklahoma | 16.1 | 6.31 | 0.23 | 6.54 | 71.9 |
| Arkansas | 17.4 | 6.88 | 0.12 | 7 | 71.2 |
| Tennessee | 16.7 | 6.4 | 0.19 | 6.59 | 69.7 |
| Kentucky | 16.8 | 6.48 | 0.14 | 6.62 | 69.8 |
| Ohio | 17.5 | 6.85 | 0.37 | 7.22 | 69.2 |
| Pennsylvania | 18.7 | 7.5 | 0.48 | 7.98 | 67.7 |
| New<br>Hampshire | 18.6 | 6.67 | 0.6 | 7.27 | 66.5 |
| Connecticut | 17.7 | 6.53 | 1.03 | 7.56 | 61.5 |
| Virginia | 20.5 | 5.7 | 0.62 | 6.32 | 66.2 |
| North<br>Carolina | 16.7 | 6.2 | 0.3 | 6.5 | 68.2 |
| South<br>Carolina | 18.2 | 6.5 | 0.32 | 6.82 | 66.7 |
| Georgia | 14.3 | 4.88 | 0.44 | 5.32 | 66.4 |
| Florida | 20.9 | 7.12 | 2.06 | 9.18 | 65.7 |
| Alabama | 17.3 | 6.7 | 0.18 | 6.88 | 68.6 |
| Mississippi | 16.4 | 6.25 | 0.12 | 6.37 | 65.8 |
| Louisiana | 15.9 | 5.93 | 0.21 | 6.14 | 69.3 |
| Texas | 12.9 | 4.19 | 0.77 | 4.96 | 67.2 |
| Nevada | 16.1 | 4.8 | 1.2 | 6 | 66.4 |
| Idaho | 16.2 | 6.05 | 0.28 | 6.33 | 67.8 |
| Wyoming | 17.1 | 6.3 | 0.12 | 6.42 | 61.5 |
| West<br>Virginia | 19.7 | 7.95 | 0.135 | 8.085 | 70 |

The correlation of the integer dividing the incidence of COVID-19 by the State population density compared to the case-fatality ratio was also carried out. The incidence of COVID-19 infection when factored with State population density as a denominator, significantly correlated (R=0.67 p<0001) with the case-fatality ratio. This latter correlation also applied for both the States with partial smoking bans (R=0.57 p<0.003) and more strict regulation (R=0.75 p<0.0001) of tobacco smoking (Tables 3. and 4.).

## DISCUSSION

The incidence of respiratory infection increases with tobacco smoking. Moreover tobacco smoking exacerbates the severity of pulmonary infections. The risk for mortality from tuberculosis has been shown to increase nine-fold in tobacco smokers (Chi-Panf et al 2010). Legionella infection is known to increase up to 121% increased risk of legionella pneumonia with every packet of cigarettes smoked (Almirall et al 2013). Similarly Mycoplasma and viral infections such as the influenza are more common in tobacco smokers (Doolittle and Davis 2018). Smokers have also been noted to have had higher mortality in the 2012 MERS-CoV outbreak (Vardavas and Nikitara 2020). In a similar fashion COVID-19 is a highly infectious viral disease that affects the respiratory system leading to a myriad of presentations and complications.

Smoking impairs pulmonary defences against lung infection(Thimmulappa et al 2012), and may be implicated in the transmission of COVID-19. The immune system is adversely affected by the components of tobacco smoking including particulate matter element. Tobacco smokers through their smoking habits are exposed to very high concentrations of particulate matter (Loffredo et al 2016). The exposure to polluted air in London is equivalent to smoking 150 cigarettes per year (British Heart Foundation 2019).

The impact of particulate matter PM2.5 on the immune response influences macrophage function and the modulation of the cytokine response. PM2.5-induced inflammation may result in an increase in the number of pulmonary neutrophils, eosinophils, T cells and mastocytes (Sigaud et al 2007) (Gripenbäck et al 2005). This cellular reaction can result in inflammatory cytokine production and the resultant cytokine storm has been responsible for a significant number of COVID-19 related deaths (Nile et al 2020).

Most of the early studies linking smoking to COVID-19 infection were carried out in mainland China. Zhou et al. assessed 191 patients infected with COVID-19, 54 of whom succumbed to the infection, while the other 137 survived resulting in a case-fatality ratio of 28.3%. The high case-fatality ratio suggests that this cohort (191) were seriously ill with COVID-19. A nonsignificant proportion (9%) of those who succumbed to the infection were currently smoking as opposed to 4% smokers among those who survived. Similar nonsignificant results were obtained by Zhang et al 2020. Out of 140 patients with COVID-19, 58 had severe infection. Of the patients with severe infection 3.4% were current smokers and 6.9% were smokers in the past, while in the less severe group none were presently smoking and only 3.7% were previous smokers. The largest study performed in China recruited 1,099 patients with COVID-19 infection (Guan et al 2020). The proportion of patients with severe symptoms comprised 15.7%. Of the patients presenting with severe symptomatology, 16.9% were recent smokers and 5.2% were ex-smokers, while in the group with minimal COVID-19 symptoms 11.8% were currently smoking and 1.3% smoked in the past.

Meta-analysis of several studies from various centres were carried out in the months following the outbreak in China. One meta-analysis analysing 11,590 hospitalized patients, indicated that which 2,133 (18.4%) developed severe disease. Disease severity occurred in 29.8% of patients who smoked currently or in the past compared with 17.6% of non smokers (OR=1.91; 95% CI: 1.42–2.59) (Patanavanich and Glantz 2020). Another meta-analysis by Simons et al. (2020) assessed 102 studies investigating the connection between disease progression and smoking status. Patients who were currently smoking were more likely to experience disease progression as opposed to non smokers (RR=1.39; 95% CI: 1.09–1.77).

This study indicated that States in the USA with high levels of tobacco smoking and partial regulation had significantly higher rates of COVID-19 infection incidences than States with more severe smoking bans. As described earlier this could be attributed to the reduced respiratory immunity due to the toxicity of tobacco smoking including particulate matter. Alternatively besides impaired pulmonary defences, transmission of COVID-19 infection through viral carriage by surface particulate matter has been alluded to by Comunian et al. Tobacco smoking has the attribute of exposing both smokers and bystanders to very high concentrations of particulate matter in a short period of time. This has been noted not only indoors with adequate ventilation, but also in the open air.

Studies have shown that average levels of PM2.5 in open-air smoking venues exceed the WHO recommendation of 25μg/m3. These levels varied from 8.32μg/m3 to 124μg/m3 at open-air settings where tobacco smoke was present. Extremely elevated levels of 1,000μg/m3 have been noted in some smoking venues. These elevated levels of PM2.5 are more likely in densely packed areas with poor ventilation, in the presence of smokers. Confirming the risks of PM2.5 exposure due to passive smoking, even smoke-free venues close to open-air smoking settings also had potentially high PM2.5 concentrations, with levels ranging from 4μg/m3 to 120.51μg/m3 (Potera 2013).

Setti et al have shown increased COVID-19 transmission has been noted was noted to coincide with elevated peaks of particulate matter (Setti et al 2020a). The same authors have suggested that the recommended social distance of 2 metres is only effective if masks are worn because particulate matter can travel up to distances of 10 metres and more (Setti et al 2020b). The importance of face protection with masks cannot be understated, as Zhang et al have shown that the use of masks was crucial in determining the disease spread in Wuhan, Italy and New York. Wearing of masks has been estimated to have reduced the number of COVID-19 infections by more than 78,000 in Italy and over 66,000 in New York City during the months of April and May 2020 (Zhang et al 2020).

In a paper linking subway particulate matter and COVID-19, it was hypothesized that the haematite-rich particulate matter may be a superior vector to surface particulate matter as it creates a microenvironment suitable for COVID-19 persistence (Muscat Baron 2020a). Whereas surface carbon-rich particulate matter has adsorbing properties on adherent substances, subway haematite-rich particulate matter may easily release any adherent COVID-19. Moreover COVID-19 has been noted to have persisted on steel objects for up to 72 hours (van Dormalen et al 2020).

Ambient salinity and the sodium chloride component in PM2.5 have been touted as protective factor against COVID-19 (Muscat Baron 2020b Muscat Baron 2020c). The particulate matter sodium chloride component has the propensity to attract water which may deter the hydrophobic C-terminal protein in the COVID-19 Spike Protein (Muscat Baron 2020c). This protective effect did not apply to the pandemic in the salinity-rich East Coast of USA. Although the states of New Jersey, Connecticut and Massachusetts have high levels of ambient salinity (Poma 2018), the incidence and case-fatality ratio was still elevated. The role of subway commuter congestion, interconnectivity and elevated subway PM2.5 levels may have eclipsed any saline related protective factors, causing high death rate in New York State (Harris 2020) and the adjacent states of Connecticut, New Jersey and Massachusetts. Similarly the exhaled tobacco derived particulate matter is devoid of any exposure to ambient salinity. In a similar manner, the protective effect of sodium chloride may not be available to particulate matter originating from exhaled tobacco fumes.

This study also indicated that population density and age beyond 75years and over was a significant factor in the case-fatality ratio. It has been shown that the case-fatality ratio varies according to the population demographics - the older the population the greater the case fatality ratio. The population in Northern Italy aged 65 years and over constituted 25% of the Lombardy population. Consequently the case-fatality ration in Italy was 9.3%. Similarly the Netherlands had a case-fatality ratio of 7.4% with a background 65+years population of 20% (Sudharsanan et al 2020). It is therefore biologically plausible that tobacco smoking in densely packed areas with elderly and vulnerable persons in the vicinity put these individuals at greater risk.

It is interesting to note that at the time of the study, Utah, the only state with a single digit percentage US smoking population had a relatively elevated COVID-19 incidence. The percentage population who smoke in Utah is 9% and its incidence was 1989/100,000 which is high for a State with a restrictive smoking regulation. The presence of neighbouring partial ban States such as Nevada, Idaho and Wyoming may explain Utah’s COVID-19 incidence. However Utah’s case-fatality ratio was the lowest in the USA, possibly due to the combined factors of low smoking prevalence, low population density and low percentage cohorts in both the 65+ (11%) and 75+ (4.4%) age groups.

The author’s home country Malta sustained two waves of the Coronavirus infection. During the first wave the incidence was low (135/100,000) and so was the case-fatality ratio (0.29%). This cannot be said for the second wave whereby both the incidence (726/100,000) and case fatality ratios (1.2%) are significantly higher. It is interesting to note that during the first wave the ban on smoking was more severe whereby smoking in public places was only allowed 10 metres away from non-smoking patrons. This ban was not applied during the second wave. Another interesting factor is that the incidence of COVID-19 infection during the second wave appeared to peter off from 106 cases/day (16^th^ September) to 21 cases/day (27^th^ September), 13 days after the first heavy rainfall (14^th^ September). This may suggest that coronavirus-laden atmospheric particulate matter could have been cleared off during the rainfall. Once the clement weather returned, the incidence of COVID-19 increased again.

There a number of limitations to the observations noted in this paper. There is some asynchrony in a number variables assessed. The population percentage of smokers was derived from 2018 data from the Centre of Disease Centre and Prevention. This also applies to the States’ population, population density and percentage of individuals deemed overweight or obese. The data regarding incidence of COVID-19 infection, case-fatality ratio and testing frequency were collected over the period between the 18^th^ and 25^th^ September so as to reduce the bias for the constantly evolving fluctuations. Testing frequency was not available for all States, however there did not appear a significant difference of the testing frequency between both groups of states in the data available. Not all the required data from the District of Columbia, Puerto Rico, American Samoa and North Mariana Islands was available for assessment, so these territories were excluded from this study. These four regions constitute 1.37% of the whole USA population. The paper did not review the inter-State differences in infrastructure, healthcare provision, lifestyle differences and the poverty index, all of which may directly or indirectly impact on infection transmission and case-fatality ratios. Moreover the State bans considered were those imposed in 2018 and since then smoking regulations may have been altered. This study presupposes the unlikely assumption that adherence to smoking bans is enacted across the USA uniformly and admittedly this is unlikely to occur as has been shown in other countries such as Israel (Baron-Epel et al 2007).

## CONCLUSION

This paper suggests that States in the USA with elevated levels of tobacco smoking and limited smoking bans had significantly higher rates of COVID-19 infection incidences than States with more severe smoking restrictions. As shown in other papers, population density and individuals aged 75 years and over demonstrated a significant correlation with the case-fatality ratio. Adjacent State smoking ban status could affect neighbouring State COVID-19 incidence. Besides the adverse effects of tobacco smoking on pulmonary defences, it would be interesting to explore the possibility of infection transmission via COVID-19 laden particulate matter from exhaled fumes derived from tobacco smoking. Second-hand smoking may be implicated in COVID-19 transmission.

## Data Availability

Data was obtained from Internet links and websites

https://coronavirus.jhu.edu/map

https://www.cdc.gov.dadastatistics

https://datausa.io

https://obs.withings.com

